# Global epidemiology, pathogenesis, immune response, diagnosis, treatment, economic and psychological impact, challenges, and future prevention of COVID-19: A Scoping review

**DOI:** 10.1101/2020.04.02.20051052

**Authors:** Brhane Berhe, Haftom Legese, Hagos Degefa, Gebre Adhanom, Aderajew Gebrewahd, Fitsum Mardu, Kebede Tesfay, Miglas Welay, Hadush Negash

## Abstract

**Background:** Globally, the novel coronavirus, is a public health problem causing respiratory infections. Since the outbreak of severe acute and Middle East respiratory syndromes coronavirus was not reported to cause human infections. Now, it become an epidemic proportion with growing number of cases and deaths.

**Methods:** A scoping review was conducted following the methodological framework. In this scoping review, 50 records published before 28 March, 2020 were included and discussed to better understand the current epidemiology, pathogenesis, immunological response, diagnosis, evasion mechanisms and suggested strategies to boost immune system, challenges, treatment, and future preventions of the virus. PubMed, BioRxiv, MedRxiv, Global Health and google scholars were searched comprehensively for articles, preprints, grey literatures, reports, websites, conference proceedings and expert information. Studies conducted in human and published in English language were included in the analysis. All the findings and statements of the review regarding the outbreak are based on published information as listed in citations.

**Results:** We identified 360 records, of which 50 studies met the inclusion criteria. We synthesized the data from the included records and dig out the deep insights of them and pooled into this review. The burden of the outbreak is worsening due to overcrowding, presence of asymptomatic carriers, scarcity of test kits, the immune escaping ability of the virus and lack of awareness.

**Conclusions and recommendations:** Due to the fast-spreading nature of COVID-19 the prevention and control strategies become challenging. It is imposing social, psychological, and socio-economic impacts. We recommended that following social distancing, isolation suspects, using personal protective equipment, health education and introducing hand-washing practices, avoiding contact with animals, vaccine development and treatment for controlling and prevention.

## Introduction

Globally, the novel coronavirus is (COVID-19), is an emerging pandemic disease causing respiratory syndrome that needs urgent attention^1^. It was not reported to cause human disease until the outbreak of severe acute respiratory syndrome (SARS-CoV) occurred early in 2003, which infected about 8000 persons, with a case fatality of 9.5%^2^, Followed by Middle East respiratory syndrome (MERS-CoV) from 2012-2015, which infected about 2500 persons worldwide and a case fatality rate of 35%^2,3^. COVID-19, is a new strain that has not been identified among humans^4^ and now become an epidemic proportion with growing number of cases rapidly from December 2019 to 571,678 globally, with 26,495 deaths as of 28 March 2020^5^.

COVID-19 is a viral infection in the family of coronaviridae. The family includes viruses which are a positive sense, enveloped, non-segmented RNA virus in the genus of beta coronavirus with zoonotic origin first described in 1931 whereby first isolated from humans in 1960s^1^. The novel coronavirus, previously designated 2019-nCoV, was identified as the cause of a cluster of pneumonia cases in Wuhan, China at 31 December 2019. On 31 December 2019, the first four cases of an acute respiratory syndrome with unknown etiology were reported in Wuhan City, China among people linked to a local seafood market^1, 3^. From 31 December 2019 through 3 January 2020, a total of 44 case-patients with pneumonia of unknown etiology were reported to WHO from China. Within one month the outbreak spreaded to 19 countries with a total of 11, 791 confirmed cases and 213 deaths^4, 5^.

After the declaration of COVID-19 as global public health emergency^6^, about 118 598 confirmed cases were reported from more than 100 countries^6, 7^. Currently, COVID-19 is affecting 199 countries and 2 territories named the *Diamond Princess* in Yokohama, Japan, and the Holland America’s MS Zaandam) with a total of 571,678 confirmed cases, 26,494 deaths and 142,474 recovered cases was reported as of 28 March 2020 2020^8^. As per the previous studies reports median age of COVID-19 cases was 59 years (15 to 89 years) with the majority being male^7^. The average incubation period of COVID-19 was estimated to be 4.8±2.6, ranging from 2 to 11 days^9,10^.

The outbreak has distributed throughout the globe. WHO and national guidelines have endorsed preventive strategies to the current outbreak. However, the current prevention and control strategies of COVID-19 is facing challenges. Hence, we conducted this scoping review to assess the current epidemiology, pathogenesis, immunological response, diagnosis, evasion mechanisms and suggested strategies to boost immune system, challenges, and future preventions.

## Methods

### Methodological framework

We used the following methodological framework to conduct this scoping review. (1) Identification review question (2), Developing review objectives, (3) Developing search strategy and identification of search sources, (4) Screening records and data extraction and (5) Setting eligibility criteria.

### Search strategy and searching sources

A search strategy was developed using specific key concepts in our research question: “epidemiology of COVID-19”, “burden of COVID-19”, “pathogenesis of COVID-19”, “COVID and immune system”, “diagnosis of COVID-19”, “immune invading and evasion mechanisms”, “current preventions and treatment strategies of COVID-19” and “challenges to the current strategies and future preventions”. We comprehensively searched PubMed, Cochrane, Global Health and google scholars. To avoid missing relevant studies we included preprints and grey literatures using BioRxiv, MedRxiv, World Health Organization and Centers for Disease Control and Prevention websites, conference proceedings and expert information.

### Study selection and data extraction

After the literature search, all the references were imported to Zotero. Four researchers (BB, HD, HL and HN) independently screened studies for eligibility and relevance. A fifth researcher (GA) was consulted for discrepancies. We resolved differences in opinion through discussion.

### Eligibility criteria

Articles, reports, preprints, expert opinions, and information from websites conducted among human participants, published in English language, and briefly showing the data regarding the current outbreak COVID-19 were included.

## Results and discussion

### Characteristics of the included studies and reports

Most of the records were retrieved from WHO, CDC reports, Global health, and websites. Out of the 50 included articles (**Figure 1**) more than half were reports and expert opinions.

### Three months Global burden of COVID-19 in terms of total confirmed cases and deaths

The confirmed cases of COVID-19 are increasing from time to time. In the first two months the number of cases were higher among countries in the West pacific Asian region than other regions where the lowest cases were reported in Africa. And later as of March 2020 the number of cases was observed to be exponentially increased among countries in the European, American, and Eastern Mediterranean regions (**Figure 2**).

Furthermore, the reports of death were predominant in the Western Pacific Asian region in January and February 2020. However, the reported deaths outnumbered among countries in European region by March 2020 (**Figure 3**).

### Pathogenesis and clinical syndrome of COVID 19

Individuals infected with COVID-19 presented with early symptom of high fever (39°c), headache and abnormal respiratory findings such as cough, and difficult breathing. The virus might pass through the mucous membranes, especially nasal and larynx mucosa then enters the lungs through the respiratory tract^11^. After the virus reach in the lung it spreads to peripheral blood, causing viremia. Then the virus would adhere and express to the angiotensin converting enzyme 2 (ACE2), of the organs like lungs, heart, renal, gastrointestinal tract. Patients infected with the virus have higher number of leukocytes, increased plasma pro-inflammatory cytokines^12-14^. The main pathogenesis of COVID-19 infection as a respiratory system targeting virus was severe pneumonia, viremia, combined with the incidence of ground-glass opacities, and acute cardiac injury^15^.

### Host immune response to COVID-19

The immune system is responsible for the controlling, resolution and immunopathogenesis of CoV infections. The immune response is due to receptor recognition between the pathogen-associated molecular patterns (PAMPs) and the pattern recognition receptors (PRRs). Usually, Toll-like receptor (TLR) 3, TLR7, TLR8, and TLR9 sense viral RNA and DNA in the endosome^16, 17^. The most important recognition mechanisms of viral RNA are viral RNA receptor (retinoic-acid inducible gene I), cytosolic receptor (melanoma differentiation-associated gene 5) and nucleotide transferase cyclic GMP-AMP synthase^18, 19^. This complex signaling recruit adaptors, including TLR-domain-containing adaptor protein, mitochondrial antiviral-signaling protein^20^ and stimulator of interferon genes protein^21^ to trigger downstream cascade molecules. This will also be involved in adaptor molecule MyD88 and lead to the activation of the transcription factor nuclear factor-κB and interferon regulatory factor 3 and the production of type I Interferons and a series of pro-inflammatory cytokines^22^. Hence, virus-cell interactions produce a diverse set of immune mediators against the invading virus^23^.

### Innate immunity

To mount an antiviral response, innate immune cells need to recognize the invasion of the COVID-19. The recognition is through PAMPs and PRRs. In the innate immunity there is activation and regulation of immune system to eliminate the virus, otherwise results in immunopathology. A few plasma cytokines and chemokines like IL-1, IL-2, IL- 4, IL-7, IL-10, IL-12, IL-13, IL-17, GCSF, macrophage colony-stimulating factor, IP-10, MCP-1, MIP-1α, hepatocyte growth factor, IFN-γ and TNF-α were observed higher among COVID-19 infected individuals^24-26^. COVID-19 causes an inflammatory response in the lower airway and led to lung injury to produce cytokine storm in the body, which may be associated with the critical condition among COVID-19 infected individuals^27^.

Like SARS-CoV and MERS-CoV, early high rise in the serum levels of pro-inflammatory cytokines were observed in SARS-CoV-2^28^, suggesting a potential similar cytokine storm-mediated disease severity of COVID-19 infection^29, 30^. Effective innate immune response involves the action of interferon responses and its downstream cascade that culminates in controlling viral replication and induction of effective adaptive immune response sharing the same attachment receptor with SARS-CoV^31^. The recognition site is present in subset of lung cells called type 2 alveolar cells^14^.

### Adaptive immune response

Unlike the other corona viruses, a limited serology details of COVID-19 were reported. In a previous study, peak IgM antibodies were observed after day 9 of disease onset. additionally, sera from 5 patients of confirmed COVID-19 showed some cross-reactivity with SARS-CoV.

Furthermore, antibodies were observed to be neutralized by COVID-19 in an in vitro plaque assay, suggesting a possible successful mounting of the humoral responses^14^.

Another study reported that CD8+ T cell responses were frequently observed than CD4+ T cell response. Generally, the virus specific T cells were the central memory phenotypes with a significantly higher frequency of polyfunctional CD4+ T cells (IFNγ, TNFα, and IL-2) and CD8+ T cells (IFNγ and TNFα). Having considered few controversial issues, strong T cell response was correlated significantly with higher neutralizing antibody while more serum TH2 cytokines (IL-4, IL-5, IL-10)^32^.

### Immune Evasion Mechanisms

Current observations indicate that coronaviruses are particularly adapted to evade immune detection and dampen human immune responses. This partly explains why they tend to have a longer incubation period, 2-11 days than other viral infections^33^. The longer incubation period is probably due to their immune evasion properties. Hence the viral antigen can escape host immune detection at the early stage. The immune evasion mechanism is potentially like SARS-CoV and MERS-CoV. The other immune escaping mechanism is inhibition of innate immune responses, inhibition of interferon recognition and signaling, immune modulation including membrane or nonstructural proteins (NS4a, NS4b, NS15), Viral mutations, Immune exhaustion, Immune deviation: TH2 bias^34-36^. Furthermore, in the adaptive immune response the evasion mechanism is due to down regulation of antigen presentation via MHC class I and MHC class II. Whenever, macrophages or dendritic cells get infected with MERS-CoV, T cells activation will be markedly diminished^37^.

### Suggested strategies to boost our immune system

In the presence of controversial ideas between immune system and COVID-19, some scientist and reports suggested that taking items listed below are some supplements to trigger the immune system^38, 39^.

- **Reducing stress**: Stress have negative impact the production of lymphocytes. As stress increase the risk of viral disease increases.
- **Exercise**: Regular exercise promotes cardiovascular health, lowers blood pressure, helps control body weight, and offers protection against diseases. Exercise also improves blood circulation, allowing immune system cells to move through the body more freely and do their job more effectively. However, intensive exercise can cause inflammation in the body that boosts undesired immune response.
- **Eat a balanced diet with fruits and vegetables:** Previous evidence suggested that lack of these nutrients can alter the immune response.
- **Avoid smoking**: Smoking tobacco leads to susceptibility to infection and lower the protective nature of the immune system.
- **Get enough sleep:** Studies showed that people who get an optimal sleep (sleeping 7-8 hours for adults and up to 10 hours for children and teenagers) are less likely to be at risk of viral infections.
- **Supplements:** Such as Vitamin C and D, Zinc and Garlic may improve the function of white blood cells that fights infection.

### Transmission, laboratory diagnosis and current treatment

#### Transmission

the virus has two main transmissions, zoonotic transmission like outbreak of SARS-CoV in 2003 and MERS-CoV in 2012/2015^40^ and anthroponotic, via direct contact or through droplets spread by coughing or sneezing from an infected individual and it is more contagious than other viruses. Moreover, there is no evidence of congenital transmission for COVID-19^11^.

#### Laboratory diagnosis

The following laboratory diagnostic techniques are used to detect COVID-19^41-43^.

- **Viral nucleic acid test:** Is the routine confirmation test for COVID-19 based on detecting a unique sequence that shows presence or absence of the virus.
- **Serological testing:** Used for outbreak investigation.
- **Viral sequencing:** After the virus is detected by nucleic acid test viral sequencing is important for monitoring genome mutation.
- **Viral culture:** It is not routine test but used for further investigation.
- **Hematological test:** Is supportive test to the routine tests for screening the distribution of cells.
- **Chest CT Scan:** It aids as supportive diagnostic method to show pneumonia. This test should be considered to confirm COVID-19 when we are under investigating of this virus.
- **Blood oxygen saturation test:** This test also uncommon and not routinely applied as confirmatory test rather used as further investigation of the virus^42^.

### Current treatment

Neither an effective vaccines nor anti-viral therapeutic agents is approved to treat COVID-19. Hence, we mostly focus on supportive care. Rapid public health interventions with antibodies, anti-viral or novel vaccine strategies are highly essential. Passive antibody therapy is considered to limit COVID-19 epidemics which can recognize epitope regions in the foreign virus particle and reduce the virus replication and disease severity^44, 45^. Although there is no specific treatment, some reports recommended that using some anti-bacterial or anti-malaria and antiviral drugs are important as pre or shortly after the onset of the virus as prophylaxis to reduce infectiousness to others by reducing viral shedding in the respiratory secretions^44^. Some of the prophylactic drugs are listed below.

#### Azithromycin

This antibacterial drug is acts by down regulate inflammatory responses and reduce the excessive cytokine production associated with respiratory viral infections^45^.

#### Chloroquine

this anti malaria drug also acts by inhibition of viral enzymes or processes such as viral DNA and RNA polymerase, viral protein glycosylation, virus assembly, new virus particle transport, and virus release. It also involves ACE2 cellular receptor inhibition, acidification at the surface of the cell membrane inhibiting fusion of the virus, and immunomodulation of cytokine release.

#### Lopinavir and Ritonavir

these ant viral drugs are acts by bind to Mpro, a key enzyme for coronavirus replication^46^.

#### Alpha interferon

as immunomodulation as adjuvant treatment^47^.

#### Acetaminophen

it considered as temperature control^48^.

#### Serum therapy

The use of monoclonal antibodies with serum therapy and intravenous immunoglobulins preparations as passive immunization^46, 49^. This can be achieved by using peptide fusion inhibitors, anti SARS-CoV-2 neutralizing antibodies, anti-angiotensin converting enzyme 2 (ACE-2) and protease inhibitors. The spike protein present on the viral membrane plays a vital role in virus entry and is the principal antigenic component responsible for inducing host immune response^47-50^.

### Economical and psychological impact of COVID 19

Following the index case of COVID-19 infected individual in China by December 2019 it started to spread to the rest of the world. It is then declared as pandemic outbreak by WHO. Since the declaration of the outbreak it leads to several economical and psychological problems in the world. Some of the impacts include disruption of the global chain supply due to the closing their boarder, slowdown of the investment, loss of revenue due to debt, increment in health spending cost, shortage of food and drugs, decrement of business travel and tightening domestic financial markets^51, 52^. Besides this, the outbreak of the disease leads to psychological trauma like fears, sadness, anxiety, and depression of the people on how to manage the disease during the hard time^53^.

### Risk groups

Individuals with Obesity, Cardiovascular disease, Respiratory diseases like Chronic obstructive pulmonary disease (COPD), asthma, bronchitis, sinusitis, Seasonal allergies/Toxic Mold exposure, Dysbiosis, Auto-immune diseases, Fibromyalgia, Neuro- degenerative disease, Cancer, Fatty Liver, Biotoxin illness and diabetes are highly susceptible to COVID-19 due to depleted cell membranes^54^.

To minimize the risk factors reports suggested that cleaning and disinfecting surfaces in home and in areas that people touch the most is effective. Following the same preventive measures as people went out. Contact healthcare providers whenever developed signs and symptoms of COVID-19. Furthermore, individuals are requested to limit shared spaces, when having guests and keeping recommended social distancing^55^.

### Challenges and future prevention of COVID 19

#### Challenges

The challenges for the effective controlling of COVID-19 outbreak include absence validated vaccine and treatment^56,57^, ability of the viral antigen to stay longer in the air, socio- cultural behaviour of people, lack of awareness, viral capacity of staying in animate objective for weeks^58^, overcrowding environment, having asymptomatic carriers, unavailability of test kits^61^, having wide host range^57^, lack of readiness to follow the recommended social distancing, difficulty of interpreting social distancing, unknown infective dose, unknown degree of infectivity prior the onset of clinical manifestation and after recovery.

### Future preventions and recommendations

#### At government level

International, National, regional governments should participate by allocating budget for training, isolation of suspects, testing and supportive cares and awareness creation.

#### At health institutions

Health institutions should also screen and early detection, giving supportive care and treatment, distributing medical protective equipment, give health education and introducing hand washing practices to customers and preparing isolation rooms^60^.

#### At community level

Creating community awareness on the transmission and early prevention, active case detection, distribution and preparing hand washing jars, utilization of hand sanitizers and respirators^61^, avoiding over-crowding^60^, avoiding intimate contact with animals^59^ and applying hand glove to protect touching different contaminates^60^ should be practiced.

#### At churches and University levels

Minimizing conferences and Sunday schools, avoiding movement along different places, avoid lecturing in classes and replace with alternative lecture methods, posting different posters that thought about the outbreak, preparing and distributing hand washing jars for the university community at the entry and exit and empowering the community on the usefulness of social distancing and on hand washing practices.

#### For upcoming researchers

As per the fast spreading outbreak of COVID-19 infection researchers should develop validated vaccine and treatment. If in case delay unenviable, substances that boost the immune response is recommended.

## Data Availability

All data are incorporated into the manuscript.

## List of abbreviations

ACE: Angiotensin converting enzyme
CD: Cluster of differentiation
CDC: Center of disease Control
COPD: Chronic obstructive pulmonary disease
COVID-19: Novel corona virus-19
MERS-CoV: Middle East respiratory syndrome
PAMPs: Pathogen associated molecular patterns
PRISMA: Preferred Reporting Items for Systematic Reviews and Meta-analyses Protocol
PRR: Pattern recognition receptors
RNA: Ribonucleic acid
SARS-CoV: Severe acute respiratory syndrome
WHO: World health organization
TH: T helper cells
TLR: Toll- like receptor

## Declarations

### Ethics approval and consent to participate

Not applicable

### Consent for publication

Not applicable

### Availability of data and material

All data are incorporated in the manuscript.

### Authors’ contributions

All authors contributed equally to conduct this review, read, and approved the final manuscript.

## Acknowledgements

Not applicable

